# MARLA: An Autonomous Agent for Medical AI Research and Development

**DOI:** 10.64898/2026.08.21.26361049

**Authors:** Xueyu Chen, Xinyang Jiang, Zifan Song, Caihua Shan, Zilong Wang, Dongsheng Li, Cairong Zhao

## Abstract

Medical AI models have made a great impact on biomedical research and real-world clinical applications, but conducting interdisciplinary medical AI research remains challenging, requiring close collaboration between clinicians and AI experts. Recent advances in large language models (LLMs) and autonomous code agents present an opportunity for low cost medical AI development, where clinicians can build AI tailored to their own research questions, even without continuous support from dedicated AI experts. However, enabling code agents to autonomously tackle complex multimodal medical AI development tasks requires clinicians to construct and supervise an AI research loop with detailed technical specifics, demanding substantial expertise in AI and computer science that they often lack. To address this challenge, we introduce the Medical AI Research Loop Agent (MARLA), an agentic framework that completely abstracts the construction and supervision of medical AI research loops from clinicians. Given a clinician-defined research intent, MARLA automatically translates high-level research goals into executable hierarchical research loops, decomposes them into verifiable sub-loops, and specifies the models, datasets, tools, and evaluation protocols required for each task. During execution, MARLA coordinates specialized code agents, monitors progress, diagnoses failures, and iteratively refines research strategies based on experimental feedback to drive the research process toward optimal outcomes. We evaluate MARLA on multimodal medical AI tasks that require closed-loop conversion from high-level clinical study objectives to trained and validated AI models. The results demonstrate MARLA’s ability to autonomously conduct complex medical AI research while substantially reducing the need for AI expertise.

## 1 Introduction

Medical artificial intelligence has emerged as a powerful tool for advancing both biomedical research and clinical practice [1]. By integrating diverse data modalities, including medical imaging, clinical records, pathology slides, and omics profiles, AI models can uncover complex biological patterns, support clinical decision-making, and accelerate scientific discovery [2, 3, 4]. Despite this promise, developing multimodal medical AI systems remains challenging, often requiring close collaboration between clinicians and AI experts to bridge domain knowledge and technical implementation. While clinicians and biomedical researchers contribute clinically meaningful questions, domain expertise, and relevant datasets, they often lack the technical expertise required to develop AI models from these assets [5, 6, 7, 8]. Conversely, AI experts can build and optimize sophisticated models but depend on close interaction with domain experts to understand clinical objectives, data constraints, and evaluation requirements [9, 10, 11]. As a result, multimodal medical AI development remains heavily dependent on interdisciplinary collaboration with substantial expertise, resources, and iterative development efforts, creating a significant barrier to the broader adoption of AI-driven research [12, 13].

Recent advances in large language models (LLMs) and code agents present a new opportunity for clinicians to develop medical AI solutions tailored to their own research questions, even without continuous support from dedicated AI experts [14, 15, 16]. By integrating reasoning, code generation, and tool use, modern code agents can autonomously execute software engineering workflows. One of the core techniques that enable code agents to tackle complex tasks is to construct iterative development loops, in which agents repeatedly generate and execute code, evaluate outcomes, refine their strategies based on feedback, and continue this loop until the objective is achieved. Extending this paradigm to medical AI suggests a new possibility: a medical AI research loop that iteratively develops, evaluates, and optimizes AI models to address clinicians’ high-level research goals.

However, realizing a fully autonomous medical AI research loop for clinicians without AI expertise remains a significant challenge, particularly for complex multimodal problems [17]. Constructing an effective research loop requires translating high-level clinical objectives into technical specifications, such as model architectures, optimization and loss function designs, and verification criteria. While clinicians can define meaningful research questions, they often lack the technical expertise to specify the exact components needed. Furthermore, executing complex multimodal research loops requires continuous expert supervision to monitor progress, identify anomalies, diagnose failures, and decide when and how to intervene, capabilities often beyond the expertise of clinicians [18, 19].

To enable clinicians to independently conduct medical AI research, we propose Medical AI Research Loop Agent (MARLA), an AI agent designed to abstract away the complexity of constructing and supervising medical AI research loops. Given a clinician’s high-level research intent, MARLA automatically constructs research loops with detailed specifications and oversees their execution by monitoring and coordinating the participating code agents.

Developing multimodal medical AI systems is inherently complex, requiring the integration of heterogeneous, high-dimensional data and the coordinated training of multiple AI models [20, 21]. To tackle this challenge, MARLA constructs a hierarchical medical research loop, where a high-level outer loop is decomposed into multiple verifiable sub-loops. Each sub-loop is responsible for a specific task and technical exploration direction, enabling complex research objectives to be pursued in a structured and manageable manner. MARLA models the dependencies among decomposed tasks and sub-loops as a directed acyclic graph (DAG), enabling downstream tasks to inherit findings, models, and development knowledge from upstream tasks, such as reusing unimodal models trained in upstream sub-loops for subsequent multimodal development. When executing the constructed loops, MARLA acts as the central supervisory agent, coordinating code agents, monitoring progress, diagnosing failures, and adaptively refining or reinitiating sub-loops with new strategies. This enables continuous improvement of the hierarchical research loop and drives the research process toward optimal outcomes. To continuously advance its research capability across various research projects, MARLA possesses a self-evolving ability that distills reusable knowledge from traces in all sub-loops and verification results into a harness of memories, Skills, tools, and code templates, allowing accumulated experience to guide future research loops [22, 23].

We evaluate MARLA on multimodal medical AI tasks that require transforming study protocols into trained and evaluated models. The experiments examine whether closed-loop, experience-guided pipeline construction can reduce manual engineering burden and support reliable medical AI development from clinician-specified research intent.

## 2 Related Work

Recent advances in LLMs have led to increasingly close collaboration between humans and AI systems. Correspondingly, the way humans interact with AI has evolved substantially. Early LLMs were primarily treated as conversational tools that generated responses to user queries, and much of the research focused on improving the utilization of information and knowledge through prompt engineering and context engineering [24, 25, 26, 27]. As model capabilities continued to improve, researchers began applying LLMs to more complex domains, including software development, data analysis, and scientific research [28]. Unlike traditional question answering, these tasks often require iterative loops of planning, execution, observation, and refinement rather than a single interaction. To address this challenge, extensive efforts have explored tool use, code agents, and harness engineering, enabling LLMs to autonomously execute complex task workflows [29, 30, 19]. Building upon these capabilities, increasing attention has been devoted to applying agents to software engineering, machine learning development, and scientific discovery, giving rise to automated research and development systems such as AutoML-GPT [31], AutoML-Agent [32], DS-Agent [33], R&D-Agent [16]. Despite differences in implementation, these systems share the common goal of reducing reliance on expert knowledge and manual intervention in complex R&D workflows.

In the medical AI domain, researchers have similarly begun exploring the use of LLMs and autonomous agents to automate medical AI research and development [34, 35]. Recent studies have introduced planning, code generation, and tool-use capabilities into biomedical research settings, advancing the automation of data analysis, model development, and scientific discovery [36, 37]. Some efforts focus on biomedical scientific discovery, leveraging multi-agent collaboration for literature analysis, hypothesis generation, data exploration, and experimental investigation [38, 39]. Other approaches aim to automate medical AI development by employing LLMs to perform data processing, model construction, hyperparameter optimization, and experimental evaluation [40]. Representative systems such as STELLA [41] and BioMedAgent [42] further integrate agentic capabilities with biomedical domain knowledge, demonstrating the potential of autonomous systems for biomedical research and medical AI development.

Despite these advances, existing systems largely focus on automating predefined tasks, such as medical question answering, biomedical knowledge discovery, data analysis, or model development [43, 44, 45]. Even in agent systems designed for biomedical research, complex research problems are typically formulated as predefined tasks and solved directly by the agents [46]. In contrast, MARLA focuses not on the autonomous solution of individual medical tasks, but on the autonomous construction and management of the entire medical AI research lifecycle. By taking a clinician-defined research intent as input, MARLA enables agents to autonomously drive the end-to-end research process, from research planning and task decomposition to model development, validation, and iterative refinement.

## 3 Method

### 3.1 Preliminary

We formulate end-to-end medical AI research as an agentic process that transforms high-level clinical objectives into executable medical AI research loops. Theoretically, each medical AI research environment can be modeled as a tuple Ω = (*O, D*, *T*, *F, K*), where *O* represents the clinician-defined research objective, D denotes the available clinical data and modalities, *T* defines the space of possible medical AI research loops, F denotes the feedback collected during loop execution, and K represents reusable research knowledge accumulated from previous development trajectories. Within this formulation, *T* ∈ *T* denotes a specific research loop constructed from the study specification, whereas *T*\* denotes the resulting loop after supervision and iterative refinement based on execution feedback. The research system is expected to achieve: (1) requirement intake, *O* × *D* × *K* → *S*, which translates the clinical objective and available data into a structured study specification; (2) loop construction, *S* × *K* → *T*, where *T* ∈ *T*, which organizes the study into a hierarchical research loop with task dependencies and parallel exploration directions; (3) loop supervision, *T* × *F* × *K* → *T*\*, which monitors and progressively refines the research process based on execution feedback; and (4) knowledge evolution, *K* × *T*\* × *F* → *K*\*, which transforms completed development trajectories into reusable research knowledge. Given a clinician-defined objective *O* and available data *D*, MARLA aims to construct and supervise an executable medical AI research loop *T*\* while continually evolving the reusable knowledge base from *K* to *K*\* through the agentic framework detailed in the subsequent sections.

As illustrated in Figure 1, MARLA realizes this process through four interconnected components. Agentic Requirement Intake derives a structured study specification from the clinician-defined research objective, available data, and relevant prior knowledge. MARLA then constructs a hierarchical medical AI research loop by decomposing the overall objective into task-specific loops, modeling their dependencies through a DAG, and instantiating parallel exploration sub-loops. During execution, MARLA continuously monitors active research loops, diagnoses execution issues from accumulated feedback, and coordinates corrective actions with Code Agents. Finally, MARLA distills development artifacts, execution traces, experimental results, and supervisory experience into reusable Skills that support subsequent medical AI research loops.

**Figure 1:**
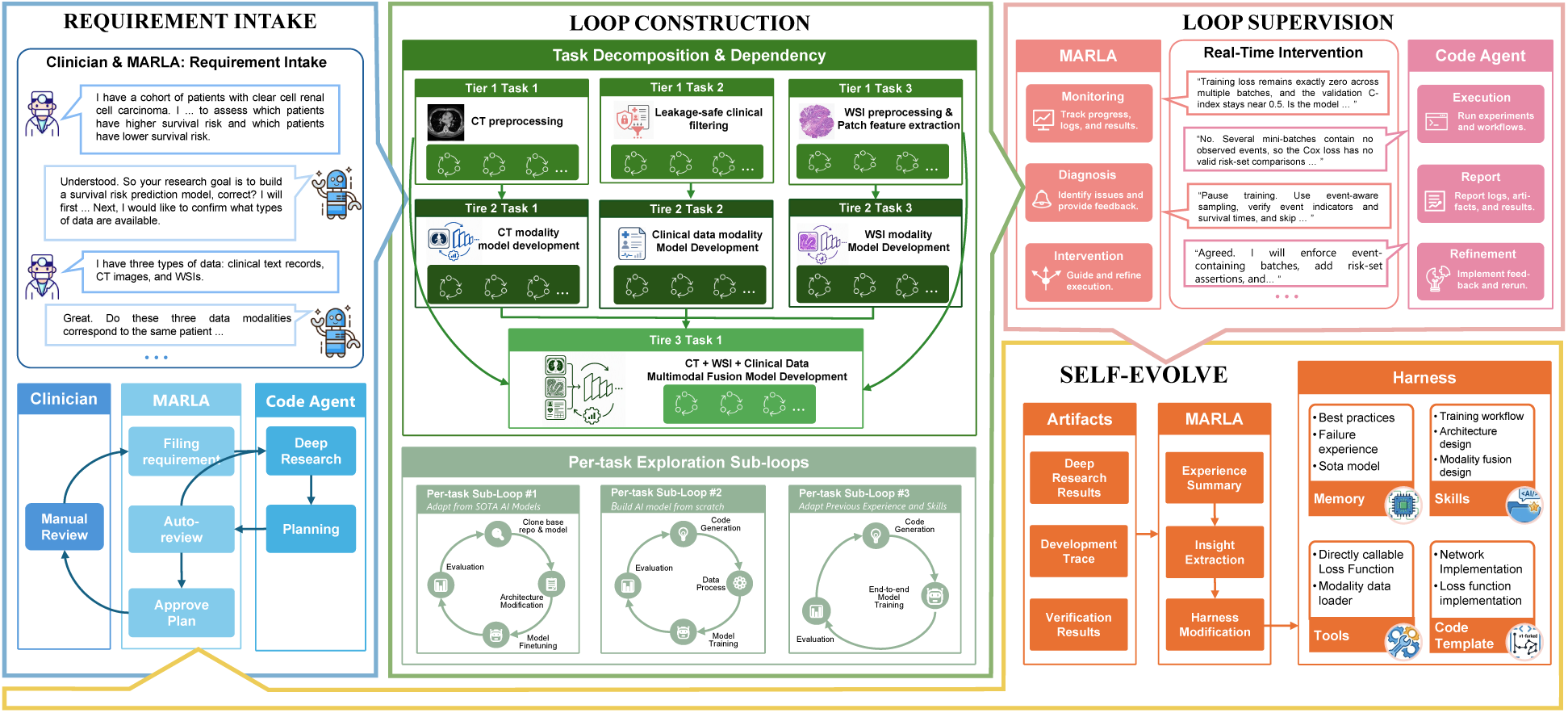
Overview of MARLA. Given a clinician-defined research intent, MARLA first conducts Agentic Requirement Intake to derive a structured specification of the target study. MARLA then constructs a hierarchical medical AI research loop by decomposing the overall objective into multiple task-specific loops, modeling their dependencies through a DAG, and instantiating parallel exploration sub-loops to investigate alternative technical directions. During execution, MARLA acts as a central supervisory agent that monitors progress, diagnoses failures, and coordinates corrective actions with Code Agents. Finally, MARLA distills execution traces, verification results, and implementation experience into reusable memories, Skills, tools, and code templates that support future medical AI research loops.

Within this framework, MARLA and Code Agents assume complementary responsibilities. MARLA performs requirement interpretation, research-loop construction, execution coordination, and supervisory intervention, whereas Code Agents carry out the underlying implementation, experimentation, and evaluation. The overall system leverages three fundamental capabilities: semantic understanding *ϕ*, code programming *ψ*, and logical reasoning *γ*. MARLA primarily employs *ϕ* and *γ* to interpret clinical intent, organize research processes, and make supervisory decisions, while Code Agents employ *ψ* to implement and evaluate candidate solutions. The following sections describe the four components in detail.

### 3.2 Agentic Requirement Intake

Agentic Requirement Intake transforms a clinician’s high-level research intent into a structured study specification that can be operationalized by subsequent medical AI research loops. Given a clinical objective *O* and available data *D*, MARLA interacts with the clinician to clarify the research question, available modalities, prediction or analysis target, evaluation requirements, and relevant clinical constraints. The clinician provides the domain knowledge required to define the study, while MARLA translates these clinical requirements into corresponding modeling, optimization, and implementation requirements. Formally, MARLA employs semantic understanding *ϕ* to interpret the clinical objective and available data, and logical reasoning *γ* to formulate a technically feasible study specification using relevant knowledge retrieved from *K*, expressed as *S* = *γ ϕ*(*O*, *D*), *K*. Here, *S* specifies the research goal, available data and modalities, prediction or analysis target, model development requirements, evaluation protocol, and expected outputs. MARLA further assesses the technical feasibility of the proposed study and identifies the modeling and experimental requirements needed to realize the clinical objective. The resulting specification is iteratively reviewed against the original clinical intent and revised whenever the objective is underspecified, inconsistent with the available data, or technically infeasible. The clinician reviews the specification from a clinical perspective, while MARLA resolves the associated AI engineering requirements. This interaction continues until the clinical objective and the technical specification are sufficiently aligned, producing an approved study specification that provides the basis for constructing the hierarchical medical AI research loop described in the following section.

### 3.3 Medical AI Research Loop Construction

Given the study specification *S*, MARLA constructs a hierarchical medical AI research loop by decomposing the overall research objective into task-specific loops, modeling their dependencies, and instantiating parallel exploration sub-loops. This construction process can be formulated as

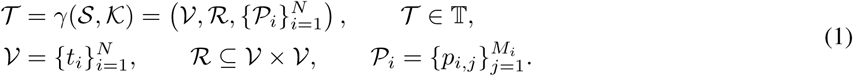

Here, *γ* denotes the logical reasoning process through which MARLA transforms the study specification *S* into a hierarchical research loop *T* using relevant knowledge from *K*. The resulting loop comprises a set of task-specific research loops *V*, their dependency relations *R*, and a collection of parallel exploration sub-loops *P_i_* associated with each task *t_i_*. Relevant knowledge retrieved from *K* informs task decomposition, dependency modeling, and the initialization of technical exploration directions.

#### Task Decomposition

A clinician-defined research objective in multimodal medical AI commonly involves multiple interdependent modeling problems that cannot be effectively addressed through a single development process. MARLA therefore decomposes the study specification S into a set of task-specific research loops V = {*t*_i_}^N^. Each task represents a distinct technical objective required to achieve the overall research goal, such as modality-specific data processing, representation learning, multimodal fusion, or downstream prediction. For each task *t*_i_, MARLA specifies its objective, required inputs, expected outputs, and completion conditions. These task specifications establish what must be accomplished before the resulting artifacts can support downstream research activities. By transforming a complex clinical objective into a set of task-specific research loops, MARLA enables individual technical problems to be developed and assessed independently while remaining aligned with the original study specification.

#### DAG Dependency among Tasks

The completion of a downstream task may depend on models, artifacts, or development knowledge produced by upstream tasks. MARLA represents these dependencies as a directed acyclic graph *G* = (*V*, *R*), in which the task-specific research loops form the nodes and the relations in *R* form the directed edges. Specifically, (*t_i_, t_j_*) ∈ *R* indicates that the outputs of task *t_i_* are required by task *t_j_*. This dependency structure enables outputs and development experience generated by upstream research loops to support downstream development. For example, unimodal research loops may provide encoders, preprocessing procedures, and empirical findings for subsequent multimodal model development. By organizing these dependencies explicitly, MARLA coordinates otherwise separate research loops into a progressive process in which upstream results inform and guide downstream decisions.

#### Parallel Exploration Sub-loops for Each Task

A task-specific research loop may admit multiple viable technical directions. For each task *t_i_*, MARLA therefore constructs *M_i_* parallel exploration sub-loops, denoted by 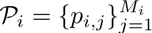, where *p_i,j_* represents the *j*-th exploration sub-loop associated with task *t*_i_. Each sub-loop pursues a distinct technical direction toward the same task objective, such as adapting an existing state-of-the-art model, developing a task-specific solution from scratch, or reusing previously acquired Skills and implementations from *K*. These parallel sub-loops operate as alternative development trajectories for the same task and produce candidate solutions, intermediate artifacts, and execution feedback. During research loop supervision, MARLA monitors their progress, examines the accumulated evidence, and provides targeted intervention when necessary. This parallel exploration strategy enables MARLA to investigate multiple potential solutions without relying on a single initial design choice.

### 3.4 Research Loop Supervision and Intervention

Once the hierarchical research loop is constructed, MARLA coordinates the execution of task-specific research loops according to their dependencies in the DAG. Root tasks without prerequisites are executed first, while downstream tasks are activated after their upstream dependencies have produced the required outputs. For each task, the corresponding exploration sub-loops operate in independent workspaces that preserve implementation files, execution traces, intermediate artifacts, and experimental results. Outputs produced by completed upstream tasks are subsequently made available to downstream tasks, allowing the research process to progressively advance from foundational development tasks toward the overall clinical objective.

Throughout execution, MARLA acts as a supervisor that continuously monitors active research loops by inspecting execution traces, intermediate outputs, and evaluation results produced by Code Agents. When anomalies, repeated failures, or unsatisfactory results are detected, MARLA analyzes the available evidence and interacts with the responsible Code Agent to identify potential causes and determine appropriate corrective actions. Depending on the diagnosis, MARLA may refine the current development strategy, provide additional guidance, restart the current sub-loop, or initiate an alternative exploration direction. Through this continual monitoring and intervention process, MARLA transforms execution feedback into actionable decisions that guide the hierarchical research loop toward more promising research outcomes. We denote the state of the *j*-th exploration sub-loop associated with task *t_i_* at iteration *r* by 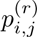, and its accumulated execution feedback by 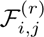. MARLA employs logical reasoning *γ*, together with relevant knowledge from K, to diagnose the current development state and determine an intervention instruction:

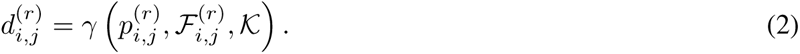

Here, 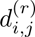 denotes the intervention instruction generated for the current sub-loop, which may involve refining the current strategy, providing additional guidance, restarting the sub-loop, or initiating an alternative exploration direction. The Code Agent subsequently employs its programming capability *ψ* to implement the intervention, advance the sub-loop, and produce updated execution feedback:

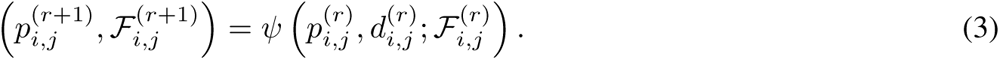

This monitoring, diagnosis, and intervention process continues until the task-specific research loop produces satisfactory outputs for subsequent research activities. Together, Eqs. 2 and 3 form a feedback-driven supervision loop in which execution feedback is continually transformed into intervention instructions and updated development states.

### 3.5 Self-Evolution through Skills

Executing a medical AI research loop produces not only final models, but also reusable procedural knowledge embedded in its development trajectories. Such knowledge includes repository-adaptation strategies, preprocessing and training procedures, tool-use patterns, and lessons from approaches that failed under particular conditions. MARLA captures this information from code modifications, execution traces, intermediate artifacts, experimental results, and supervisory decisions, and distills it into reusable Skills rather than retaining it solely as task-specific history. Each Skill describes a development objective together with the procedural guidance required to accomplish it, such as using a particular tool, adapting an existing codebase, constructing a modeling workflow, or recovering from a recurrent failure. MARLA abstracts successful trajectories into reusable procedures and unsuccessful trajectories into failure-aware lessons that record ineffective conditions and corresponding corrective actions. Given the exploration trajectories and execution feedback accumulated across the research loop, the knowledge evolution process is formulated as

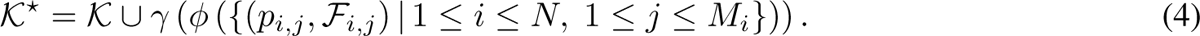

where *ϕ* extracts transferable information from the exploration trajectories and their accumulated feedback, while *γ* organizes the extracted information into reusable Skills. The resulting *K*\* represents the updated knowledge base containing experience distilled from both successful and unsuccessful development trajectories.

The accumulated Skills subsequently support different stages of medical AI research. MARLA retrieves relevant Skills to inform requirement intake, initialize exploration directions during research loop construction, guide Code Agents during development, and avoid previously observed failure modes. Skills acquired from an earlier exploration sub-loop can support later iterations of the same task, downstream task-specific research loops, and related medical AI studies. Through this continual transformation and reuse of development experience, MARLA progressively improves its ability to construct and supervise medical AI research loops beyond a single development run.

## 4 Experiments

### 4.1 Datasets

We evaluate MARLA on two publicly available multimodal medical datasets covering distinct clinical domains and prediction tasks: clear cell renal cell carcinoma survival prediction and Alzheimer’s disease diagnosis and progression prediction. To ensure fair multimodal comparison, all experiments are conducted under a modality-complete setting, where only subjects with all required modalities available are included.

#### TCGA-KIRC

The TCGA Kidney Renal Clear Cell Carcinoma (TCGA-KIRC) cohort contains multimodal data for clear cell renal cell carcinoma patients [47]. Under the modality-complete setting, we retain 237 patients with matched clinical records, CT, and whole-slide images (WSIs). The selected cohort contains 85 observed death events with a median follow-up time of 1,355 days. The clinical modality includes staging information, histopathological diagnosis, demographic variables, and treatment records, with survival-related variables removed to prevent label leakage.

#### ADNI

We evaluate our method for AD diagnosis and progression prediction on the ADNI dataset [48]. Following the modality-complete setting, we retain 695 subjects from the ADNI-1, ADNI-2 and ADNI-3 cohorts with complete clinical assessments, functional MRI (fMRI), and structural MRI (sMRI) data. The selected cohort includes 399 normal controls (NC), 228 mild cognitive impairment subjects (MCI), and 68 Alzheimer’s disease patients (AD). The clinical modality consists of demographic information, cognitive assessments, neuropsychiatric evaluations, and medical history variables (46 dimensions after leakage-prone variables are removed). The fMRI modality is represented by BOLD time series from 450 brain regions across 160 time points. The sMRI modality consists of baseline T1-weighted structural MRI scans.

### 4.2 Experimental Settings

#### Prediction tasks and evaluation metrics

For TCGA-KIRC, we evaluate overall survival prediction under single-modality, bimodal, and trimodal settings, resulting in seven survival prediction tasks, with performance measured by the concordance index (C-Index). For ADNI, we evaluate three Alzheimer’s disease classification tasks (NC vs. AD, NC vs. MCI, and sMCI vs. pMCI) under the same modality settings, yielding 18 classification tasks in total. Following prior work, classification performance is assessed using Accuracy (ACC), Sensitivity (SEN), and Specificity (SPE). All reported results are averaged over five-fold cross-validation.

#### Baselines

We compare MARLA against five representative agent systems spanning general-purpose coding agents, AutoML frameworks, scientific discovery agents, and biomedical research agents. To ensure a fair comparison, all agent-based baselines except Claude Code use GPT-5.5 as their underlying LLM backbone. MARLA is also built on GPT-5.5 and implemented using the Microsoft Copilot SDK for fully autonomous end-to-end medical AI development. Unless otherwise specified, all methods are evaluated with a budget of three candidate solutions. For methods that natively support candidate generation (e.g., MARLA and AutoML-Agent), three candidates are generated within a single run. For methods without this capability, we perform three independent runs and treat the resulting solutions as candidates under the same budget.

*Claude Code (GLM-4.7)*^1^ serves as a strong coding-agent baseline capable of code generation, debugging, experiment execution, and iterative refinement through tool use. Because Claude Code does not natively orchestrate the end-to-end medical AI development workflow, human intervention is provided solely for task coordination and workflow management.

*AutoML-Agent* [32] is a multi-agent AutoML framework that automates the machine learning lifecycle, from data preparation to model deployment, through retrieval-augmented planning and specialized agent collaboration.

*STELLA* [41] is a biomedical scientific discovery framework based on self-evolving multimodal agents that perform iterative knowledge acquisition, reasoning, and experimental exploration.

*BioMedAgent* [42] is a domain-specific biomedical research agent system that automates machine learning development through coordinated expert agents specialized in medical AI tasks.

### 4.3 Main Results

#### Survival prediction

Table 1 reports the TCGA-KIRC survival prediction results under seven modality settings. MARLA consistently achieves the best performance across all unimodal and multimodal configurations. Under unimodal settings, MARLA attains the highest C-index on Text (0.857), CT (0.726), and WSI (0.746), outperforming both domain-specific biomedical agents (e.g., BioMedAgent) and foundation model research assistants (e.g., Claude Code). These results indicate that MARLA can automatically construct effective medical AI development processes across diverse biomedical modalities without requiring manually designed modeling pipelines.

**Table 1:** Performance on TCGA-KIRC survival-risk prediction across modality settings. Results are reported as C-index (mean±std). The best result for each modality setting is shown in bold.

| Method | Uni-Modality |  |  | Multi-Modality |  |  |  |
| --- | --- | --- | --- | --- | --- | --- | --- |
|  | Text | CT | WSI | Text+CT | Text+WSI | CT+WSI | Text+CT+WSI |
| AutoML | 0.752±0.051 | 0.569±0.097 | 0.568±0.035 | 0.570±0.097 | 0.549±0.064 | 0.567±0.096 | 0.574±0.105 |
| STELLA | 0.744±0.138 | 0.606±0.058 | 0.586±0.026 | 0.558±0.071 | 0.756±0.045 | 0.583±0.088 | 0.609±0.053 |
| BioMedAgent | 0.748±0.066 | 0.609±0.073 | 0.579±0.076 | 0.664±0.072 | 0.695±0.051 | 0.539±0.065 | 0.595±0.049 |
| Claude Code | 0.626±0.136 | 0.718±0.115 | 0.701±0.053 | 0.745±0.069 | 0.737±0.041 | 0.705±0.053 | 0.701±0.100 |
| MARLA | **0.857**±0.008 | **0.726**±0.039 | **0.746**±0.077 | **0.875**±0.037 | **0.845**±0.009 | **0.744**±0.035 | **0.889**±0.033 |

The advantage becomes substantially more pronounced in multimodal settings. MARLA further improves to a C-index of 0.889 under the full Text+CT+WSI setting, surpassing the strongest baseline, Claude Code (0.701), by 0.188. Notably, increasing modality does not consistently translate into better performance for existing agent-based systems, with several baselines showing limited gains or even degradation under multimodal settings. By contrast, MARLA continues to improve as complementary modalities are incorporated, ultimately reaching a C-index of 0.889 under the full-modality configuration. This suggests that hierarchical research loop construction, cross-task knowledge inheritance, and continuous supervisory refinement enable MARLA to more effectively coordinate the development of complex multimodal models than existing agent-based research systems.

#### AD classification

Table 2 reports results on three AD-related classification tasks. While most methods achieve near-saturated performance on the relatively easier NC vs. AD task, larger differences emerge in the more challenging NC vs. MCI and sMCI vs. pMCI settings. Across both unimodal and multimodal, MARLA consistently matches or exceeds the performance of specialized biomedical research agents such as STELLA and BioMedAgent, while maintaining strong performance across all modality combinations.

**Table 2:** Performance comparison on three AD prediction tasks across ADNI dataset. C, F, and S denote clinical text, fMRI, and sMRI modalities, respectively. The best result under each modality setting is highlighted in bold.

| Modality | Method | NC vs. AD |  |  | NC vs. MCI |  |  | sMCI vs. pMCI |  |  |
| --- | --- | --- | --- | --- | --- | --- | --- | --- | --- | --- |
|  |  | ACC | SEN | SPE | ACC | SEN | SPE | ACC | SEN | SPE |
| Uni-Modality |  |  |  |  |  |  |  |  |  |  |
| C | AutoML | 0.993 | 0.985 | 0.995 | 0.871 | 0.847 | 0.885 | 0.636 | 0.522 | 0.667 |
| C | STELLA | 0.991 | 0.954 | **0.998** | 0.874 | 0.838 | 0.895 | 0.729 | 0.604 | 0.761 |
| C | BioMedAgent | 0.991 | 0.969 | 0.995 | 0.860 | 0.856 | 0.862 | 0.728 | 0.476 | 0.794 |
| C | MARLA | 0.993 | 0.986 | 0.995 | 0.857 | **0.882** | 0.842 | 0.744 | 0.749 | 0.739 |
| F | AutoML | 0.763 | 0.482 | 0.809 | 0.574 | 0.518 | 0.607 | 0.649 | 0.498 | 0.689 |
| F | STELLA | 0.750 | 0.676 | 0.762 | 0.576 | 0.491 | 0.624 | 0.548 | 0.736 | 0.500 |
| F | BioMedAgent | 0.833 | 0.399 | 0.907 | 0.592 | 0.456 | 0.669 | 0.618 | 0.569 | 0.633 |
| F | MARLA | 0.641 | 0.882 | 0.599 | 0.663 | 0.583 | 0.709 | 0.591 | 0.689 | 0.567 |
| S | AutoML | 0.735 | 0.368 | 0.797 | 0.538 | 0.584 | 0.511 | 0.776 | 0.264 | 0.911 |
| S | STELLA | 0.745 | 0.560 | 0.777 | 0.549 | 0.474 | 0.591 | 0.662 | 0.338 | 0.338 |
| S | BioMedAgent | 0.728 | 0.416 | 0.782 | 0.547 | 0.496 | 0.577 | 0.701 | 0.331 | 0.800 |
| S | MARLA | 0.792 | 0.828 | 0.787 | 0.664 | 0.724 | 0.629 | 0.711 | 0.607 | 0.739 |
| Multi-Modality |  |  |  |  |  |  |  |  |  |  |
| C+F | STELLA | 0.976 | 0.955 | 0.980 | 0.861 | 0.864 | 0.860 | 0.754 | 0.624 | 0.789 |
| C+F | BioMedAgent | 0.910 | 0.971 | 0.900 | 0.813 | 0.759 | 0.845 | 0.693 | 0.329 | 0.789 |
| C+F | MARLA | **0.998** | **1.000** | **0.998** | 0.866 | 0.838 | 0.882 | 0.732 | 0.687 | 0.744 |
| C+S | STELLA | 0.989 | **1.000** | 0.988 | 0.841 | 0.787 | 0.870 | 0.623 | 0.516 | 0.650 |
| C+S | BioMedAgent | 0.991 | 0.956 | **0.998** | 0.831 | 0.781 | 0.860 | 0.763 | 0.147 | 0.928 |
| C+S | MARLA | 0.996 | 1.000 | 0.995 | 0.867 | 0.837 | 0.884 | 0.656 | 0.667 | 0.654 |
| C+F+S | STELLA | 0.987 | 0.989 | 0.985 | 0.866 | 0.776 | **0.917** | 0.812 | 0.358 | **0.933** |
| C+F+S | BioMedAgent | 0.985 | 0.956 | 0.990 | 0.794 | 0.745 | 0.822 | 0.496 | 0.582 | 0.472 |
| C+F+S | MARLA | 0.995 | 0.994 | 0.995 | **0.873** | 0.819 | 0.904 | 0.813 | **0.793** | 0.832 |

Notably, under the full multimodal setting (C+F+S), MARLA achieves the best ACC (0.873) on NC vs. MCI and the best ACC (0.813) and SEN (0.793) on sMCI vs. pMCI. The greatest advantage is observed in more clinically challenging classification tasks, where disease progression patterns must be inferred from heterogeneous clinical and neuroimaging information. These results indicate that MARLA can effectively achieve multimodal model development and leverage cross modal complementary information to achieve powerful performance beyond existing agent-based research systems in complex classification problems.

### 4.4 Ablation Study

Table 3 presents the ablation results on TCGA-KIRC survival prediction. Overall, each component contributes to improving predictive performance or development efficiency. Among all variants, removing Agentic Requirement Intake results in the largest performance degradation, particularly under unimodal settings. Without a structured interpretation of the clinical objective and its corresponding technical requirements, Code Agents frequently pursue suboptimal development trajectories, resulting in ineffective model designs and unreliable experimental outcomes. Consequently, the C-index drops substantially to 0.542, 0.576, and 0.552 on Text, CT, and WSI, respectively. Although the degradation is less severe in multimodal settings, performance remains consistently below the full system. This suggests that accurately translating clinician-defined research intent into executable research specifications is critical for constructing effective medical AI research loops.

**Table 3:** Ablation study of MARLA on TCGA-KIRC survival prediction. We report C-index under different modality settings. Runtime denotes the estimated serial execution time required to complete all seven tasks.

| Method | Uni-Modality |  |  | Multi-Modality |  |  |  | Efficiency |
| --- | --- | --- | --- | --- | --- | --- | --- | --- |
|  | Text | CT | WSI | Text+CT | Text+WSI | CT+WSI | Text+CT+WSI | Runtime (h) |
| **MARLA** | **0.857** | **0.726** | **0.746** | **0.875** | **0.845** | **0.742** | **0.889** | 4.6 |
| **w/o Requirement Intake** | 0.542 | 0.576 | 0.552 | 0.796 | 0.780 | 0.607 | 0.629 | 3.8 |
| **w/o Skill Retrieval** | 0.847 | 0.665 | 0.560 | 0.825 | 0.831 | 0.653 | 0.873 | 4.0 |
| **w/o Web Search** | 0.850 | 0.726 | 0.702 | 0.875 | 0.838 | 0.728 | 0.884 | 4.5 |
| **w/o Parallel Exploration** | 0.850 | 0.726 | 0.645 | 0.747 | 0.846 | 0.683 | 0.853 | 25+ |

Removing Skills consistently degrades performance across both unimodal and multimodal settings, with particularly large drops on WSI (0.560) and the full multimodal setting (0.873). These results indicate that reusable development knowledge accumulated from previous research loops provides valuable guidance for subsequent model development and facilitates effective transfer of expertise across tasks and modalities. In contrast, removing Web Search leads to only minor performance changes on most tasks. This suggests that external repositories and online resources primarily serve as complementary knowledge sources, while the majority of development expertise can be acquired and retained through self-evolving Skills.

In contrast, removing parallel exploration sub-loops primarily impacts development efficiency. The estimated serial runtime increases dramatically from 4.6 hours to more than 25 hours, despite only a moderate reduction in predictive performance. In this setting, each task is restricted to a single development trajectory, eliminating MARLA’s ability to explore alternative technical directions in parallel. As a result, the system spends substantially more time iterating on suboptimal solutions before reaching a satisfactory outcome. These findings suggest that parallel exploration primarily improves development efficiency by reducing ineffective search and accelerating convergence toward promising modeling strategies.

Taken together, these results suggest that effective medical AI research automation requires accurate translation of clinical objectives into executable research specifications, continual reuse of accumulated development knowledge, access to complementary external information, and efficient exploration of alternative technical directions.

### 4.5 Robustness Across LLM Backbones

Figure 2 presents MARLA with five LLM backbones across seven modality settings. Despite differences among the backbone models, MARLA consistently performs well in both unimodal and multimodal tasks, indicating that the proposed framework is still effective across different LLM backbones. Multimodal configurations are usually superior to unimodal settings, indicating that MARLA can effectively utilize complementary information across biomedical modalities. The performance differences between the backbones are relatively moderate. GPT-5.5 achieved the highest overall C-index (0.889) under the full Text+CT+WSI setting, while Claude-Opus-4.6 achieved a comparable performance of 0.863. Even smaller models, including GPT-5.6-Sol and Claude-Connect-5, can consistently provide competitive solutions in most modality settings. Collectively, these findings suggest that MARLA’s effectiveness is not tied to a particular LLM backbone. Instead, MARLA consistently supports the construction, supervision, and iterative refinement of medical AI research loops across different backbone models, demonstrating the framework’s robustness to backbone selection.

**Figure 2:**
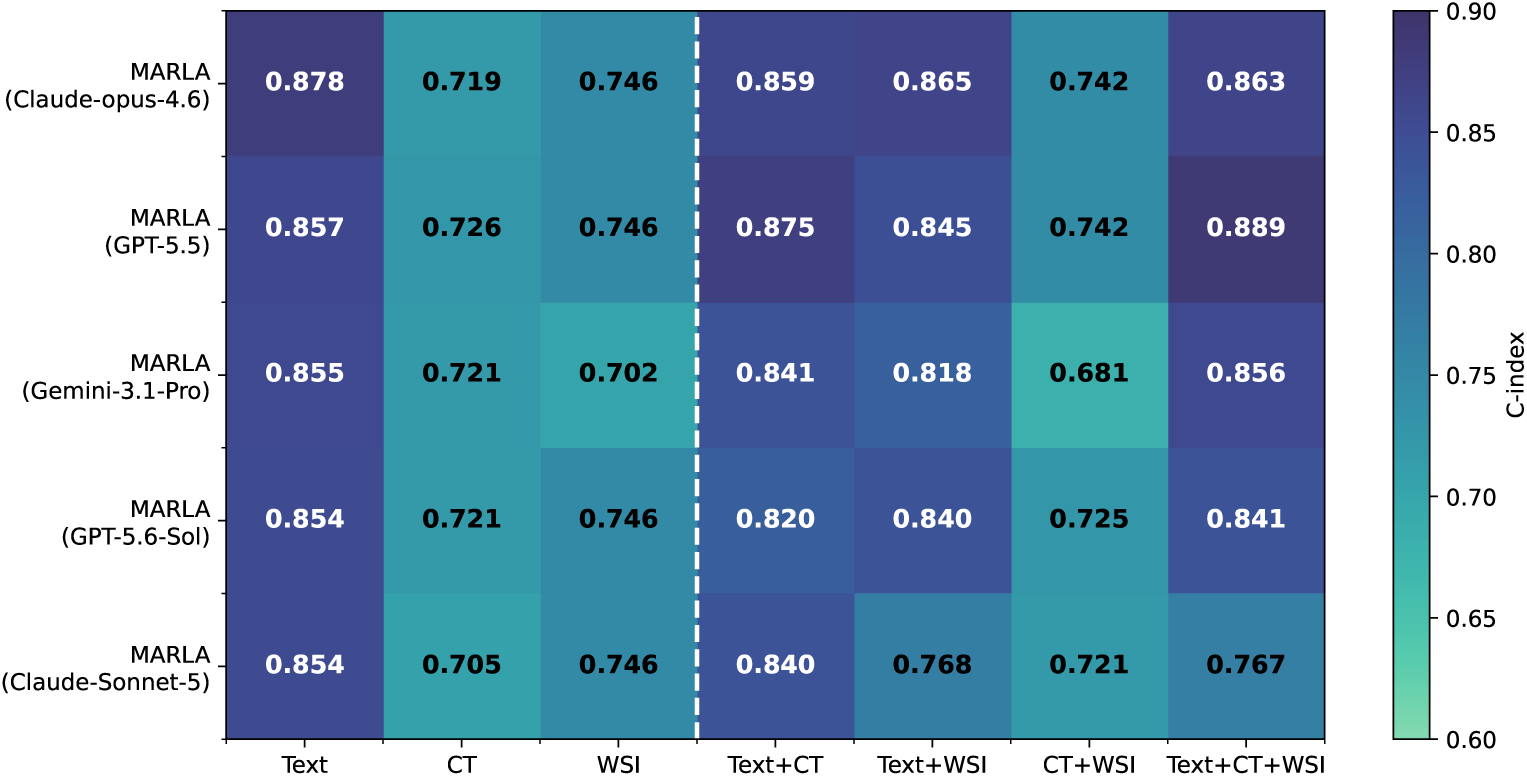
Performance of MARLA across different LLM backbones and modality configurations on TCGA-KIRC survival prediction. Results are reported as C-index. We evaluate five LLM backbones (Claude-Opus-4.6, GPT-5.5, Gemini-3.1-Pro, GPT-5.6-Sol, and Claude-Sonnet-5) under both unimodal and multimodal settings. The consistently strong performance across model families demonstrates the robustness and portability of MARLA’s research loop construction and supervision capabilities.

### 4.6 Diverse Medical AI Research Settings

TCGA-KIRC and ADNI evaluate MARLA under modality-complete settings using five-fold cross-validation. In contrast, the development of real-world healthcare artificial intelligence typically involves vastly different research environments, including modal loss, class imbalance, and predefined training/validation/testing segmentation. We therefore further evaluate MARLA on MIMIC-IV-MM [49], a four-modality healthcare prediction benchmark containing Chest X-ray (CXR), electrocardiograms (ECG), Clinical Note, and electronic health records (EHR).

Unlike previous datasets, MIMIC-IV-MM contains a large number of missing modalities, with only a small portion of samples having all four modalities. Therefore, MARLA needs to develop multimodal solutions with missing modal awareness using a complete queue, rather than limiting development to a complete subset of modalities. This benchmark further evaluates three healthcare prediction tasks, including in-hospital mortality prediction, hospital readmission prediction, and extended hospital stay prediction under pre-defined segmentation and varying degrees of class imbalance.

Table 4 reports MARLA’s performance under both unimodal and multimodal settings. Despite substantial modality missingness and heterogeneous task characteristics, MARLA consistently achieves strong performance across all three prediction tasks. MARLA further benefits from multimodal integration and achieves strong performance in various modal configurations, demonstrating robust adaptation to the challenges introduced by MIMIC-IV-MM. These findings extend beyond performance improvements alone. While TCGA-KIRC evaluates multimodal survival prediction under modality-complete settings and ADNI focuses on disease classification using five-fold cross-validation, MIMIC-IV-MM introduces additional real-world challenges, including missing modalities, severe class imbalance, and predefined train-validation-test splits. These results indicate that MARLA generalizes across diverse medical AI research settings by adapting its development and evaluation strategies to task-specific requirements rather than following a predefined workflow.

**Table 4:** Performance of MARLA on three healthcare prediction tasks from the MIMIC-IV-MM dataset under unimodal and multimodal settings. The dataset contains four healthcare modalities, including CXR, ECG, Clinical Note, and EHR, with substantial modality missingness. Models are trained using all available samples with missing-modality-aware fusion. We report Accuracy (ACC), AUROC, AUPRC, macro-average Precision (mAP), Recall (mAR), and F1 (mF1). The best results in each column are shown in bold.

| Modality | Task | ACC | AUROC | AUPRC | mAP | mAR | mF1 |
| --- | --- | --- | --- | --- | --- | --- | --- |
| EHR | Mortality | 0.8087 | 0.8595 | 0.2701 | 0.5870 | 0.6929 | 0.6007 |
|  | Readmission | 0.8500 | 0.4810 | 0.1140 | 0.5171 | 0.5190 | 0.5178 |
|  | Long Stay | 0.6729 | 0.7594 | 0.7610 | 0.6844 | 0.6760 | 0.6700 |
| Clinical Note | Mortality | 0.6979 | 0.7486 | 0.3071 | 0.5391 | 0.6080 | 0.5146 |
|  | Readmission | 0.6667 | 0.3804 | 0.0705 | 0.4738 | 0.4269 | 0.4299 |
|  | Long Stay | 0.5464 | 0.6239 | 0.5813 | 0.5522 | 0.5515 | 0.5460 |
| ECG | Mortality | 0.6707 | 0.7303 | 0.1597 | 0.5501 | 0.6689 | 0.5096 |
|  | Readmission | 0.5735 | 0.5302 | 0.1027 | 0.4982 | 0.4937 | 0.4198 |
|  | Long Stay | 0.5663 | 0.5917 | 0.6207 | 0.5748 | 0.5743 | 0.5662 |
| CXR | Mortality | 0.6957 | 0.7935 | 0.2736 | 0.5742 | **0.7332** | 0.5462 |
|  | Readmission | 0.7200 | **0.6413** | **0.1602** | **0.5239** | **0.5625** | **0.5039** |
|  | Long Stay | 0.5327 | 0.5612 | 0.5631 | 0.5341 | 0.5339 | 0.5323 |
| Multimodal | Mortality | **0.9130** | **0.8606** | **0.5072** | **0.6986** | 0.6986 | **0.6986** |
|  | Readmission | **0.8900** | 0.4416 | 0.0799 | 0.4588 | 0.4837 | 0.4709 |
|  | Long Stay | **0.7477** | **0.7787** | **0.7900** | **0.7496** | **0.7488** | **0.7476** |

### 4.7 Effectiveness of Knowledge Inheritance Across Research Loops

In order to evaluate the effectiveness of knowledge inheritance implemented by research loop dependencies, we investigated whether the experience gained through previous unimodal research loops would be beneficial for subsequent multimodal development. In order to eliminate the influence of external accumulated experience, the initial text, CT, and WSI loops were executed without prior skills, relying entirely on autonomous exploration, and three candidate solutions were developed in parallel. Then, the skills extracted from these unimodal loops are either retained or provided to downstream multimodal loops. Both settings use Claude-Opus-4.6 as the backbone and follow the same candidate budget, so that the comparison can separate experience from early unimodal loops through skills.

As shown in Figure 3, prior-loop Skills improve downstream performance across all four multimodal settings. The C-index increases by 0.011 for Text+CT, 0.005 for Text+WSI, 0.035 for CT+WSI, and 0.001 for the trimodal setting. It is worth noting that the improvement of CT+WSI is the greatest, and its performance is the lowest without previous cyclic skills. The significant increase from 0.7243 to 0.7597 indicates that previous loop skills are particularly beneficial when downstream loops face more challenging development problems and cannot easily determine effective multimodal solutions through independent exploration. Skill reuse also reduced code generation time from 23.3 minutes to 17.1 minutes, equivalent to a reduction of 26.6%. Due to the fact that the unimodal loop itself begins without prior skills, these benefits come from the experience generated during the current research process, rather than from externally accumulated solutions. Therefore, the results indicate that knowledge inherited from upstream task specific loops can provide effective program guidance for dependent downstream loops, improving multimodal development without the need for each task to explore autonomously from scratch.

**Figure 3:**
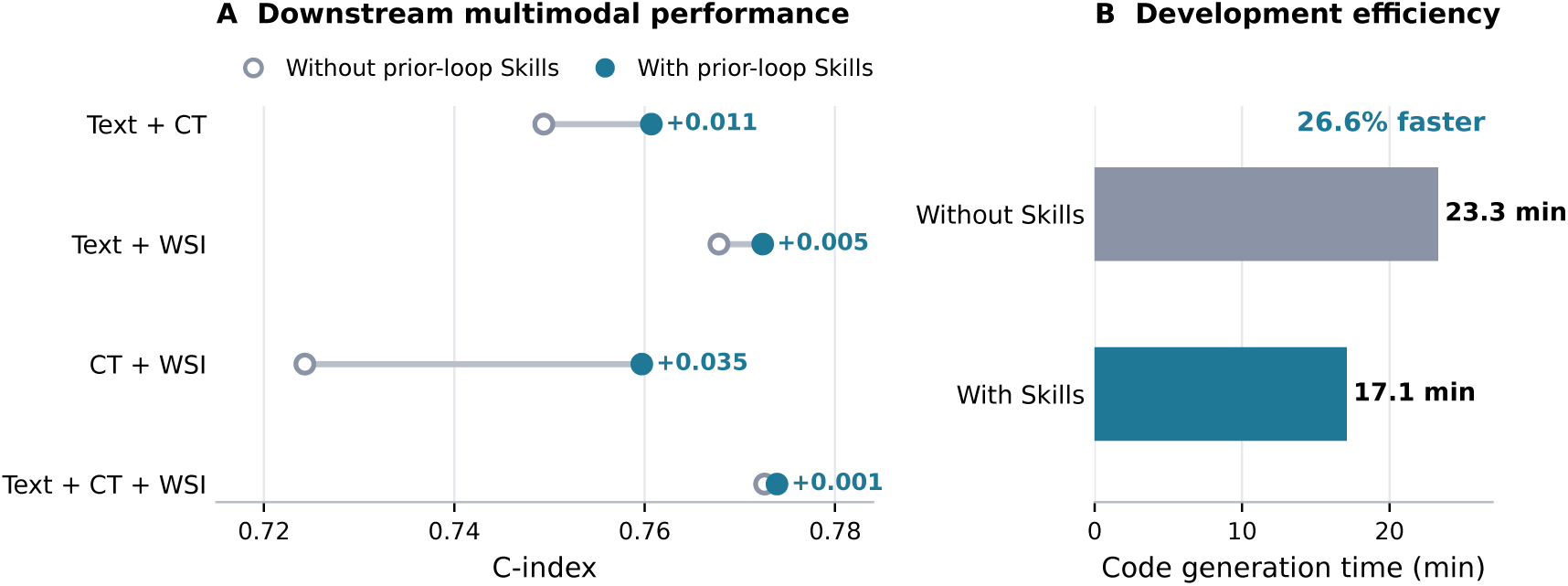
Effect of inter-loop Skill transfer on downstream multimodal development. The initial text, CT, and WSI research loops start without prior Skills and autonomously explore three candidate solutions in parallel. The Skills distilled from these unimodal loops are either withheld from or provided to subsequent multimodal loops. **(A)** C-index across four downstream multimodal settings with and without prior-loop Skills. **(B)** Code-generation time for downstream multimodal development. All experiments use Claude-Opus-4.6 as the backbone.

### 4.8 Research Loop Supervision Analysis

To better understand how MARLA supervises research loop execution, we analyze the supervisory behaviors observed during multimodal medical AI development. Figure 4 summarizes the major categories of supervision events together with representative cases of fault diagnosis and resolution. MARLA autonomously identifies a wide range of execution issues, including data integrity issues, inconsistencies between checkpoints and their associated data sources, mismatched feature shapes, resource allocation failures, and concurrency related execution errors. These observations indicate that the development of complex medical artificial intelligence requires continuous supervision, not just code generation, as many critical failures stem from experimental design, data management, and workflow coordination, rather than implementation errors.

**Figure 4:**
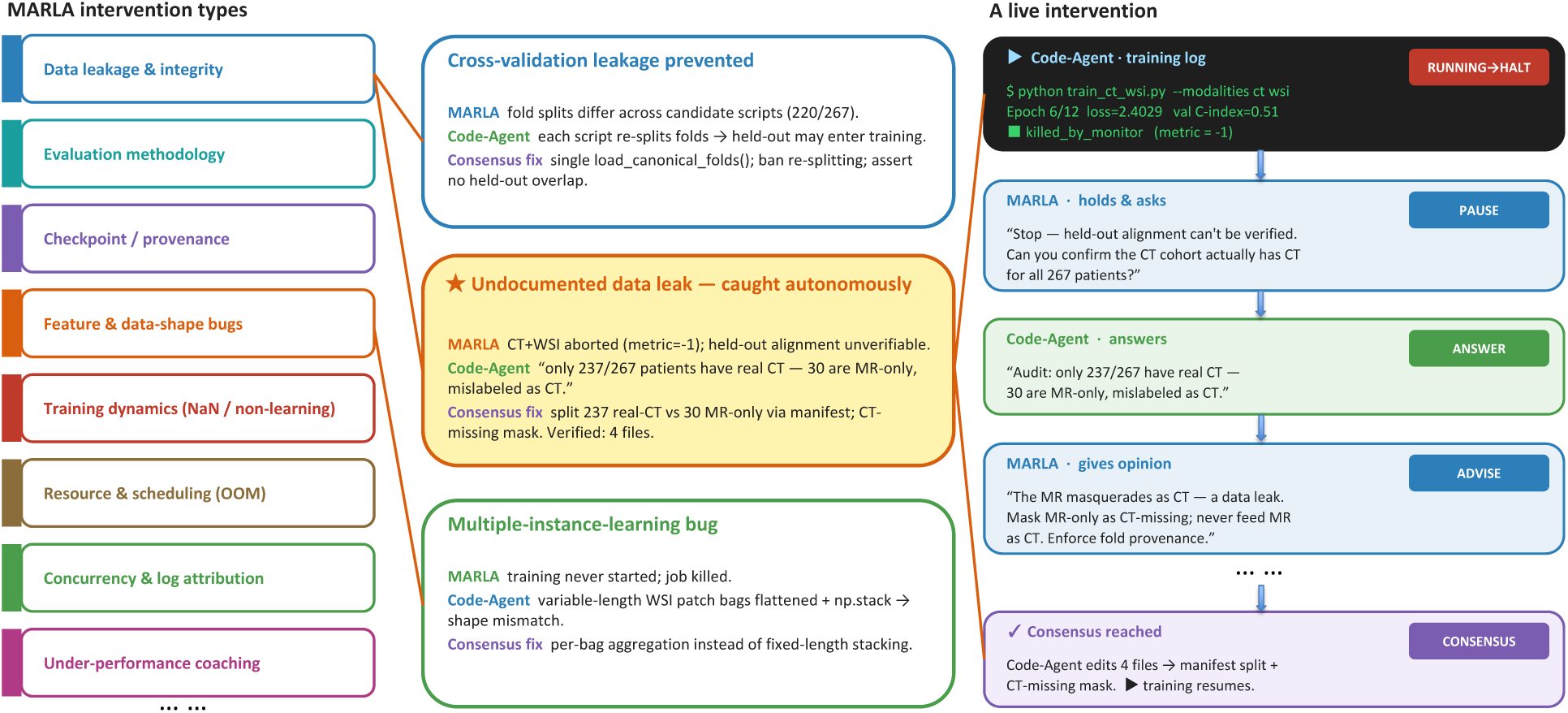
Examples of MARLA’s supervisory behaviors during research loop execution. Left: a taxonomy of supervision categories observed during multimodal medical AI development. Middle: Three representative diagnostic to solution cases involving cross validation leakage, unrecorded data leakage, and multi instance learning failure. Right: Real time MARLA - Code Proxy Supervision Process, where MARLA autonomously detects data integrity anomalies, suspends execution, collects additional evidence through interaction with the code proxy, and coordinates corrective measures before training resumes.

The middle and right panels of Figure 4 present representative cases of MARLA’s supervision. In one case, MARLA identified an inconsistency in fold assignment that could have introduced data leakage during cross-validation. In another, it detected an undocumented mismatch between execution modes that resulted in hidden data leakage. MARLA also diagnosed a multiple-instance learning failure caused by fixed-length stacking of variable-length WSI patch bags and coordinated a correction using per-bag aggregation. Rather than relying on predefined rules, MARLA collects execution evidence, interacts with the responsible code agents, evaluates alternative explanations, and coordinates corrective actions before downstream execution resumes. Together, these cases demonstrate MARLA’s role as an active supervisory agent that continuously monitors research-loop execution and guides the resolution of faults when they arise.

Figure 5 further quantifies the effects of research-loop supervision. Most supervision events involve checkpoint validation, data-integrity verification, or concurrency-related execution issues. Notably, in the CT+WSI setting, MARLA’s automatic identification and correction of data leakage increased the mean five-fold cross-validation C-index from 0.694 to 0.725. This result shows that MARLA’s supervision extends beyond fault diagnosis and can directly improve the reliability and predictive performance of downstream medical AI models.

**Figure 5:**
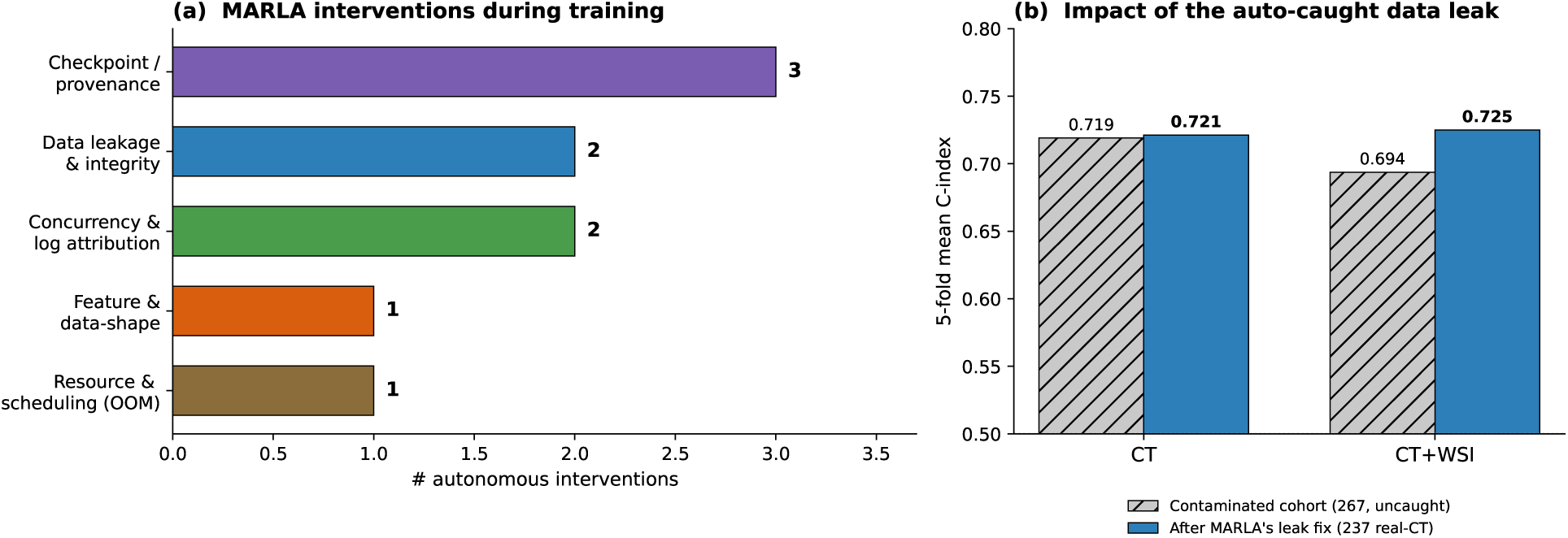
Quantitative analysis of MARLA’s research loop supervision on survival prediction tasks. (a) Distribution of supervision events observed during multimodal medical AI development. (b) Impact of autonomously correcting a detected data-leak issue on downstream survival-prediction performance. Results are reported as five-fold mean C-index.

## 5 Conclusion

Clinicians and biomedical researchers can typically define meaningful research questions and collect relevant multimodal data, but transforming these assets into effective artificial intelligence models still requires a significant amount of AI expertise and iterative development work. In this work, we introduced MARLA, a medical AI research loop agent aimed at abstracting the complexity of constructing and supervising medical AI research loops. Given the research intent defined by clinicians, MARLA will automatically construct a detailed and standardized research loop, and supervise its execution through monitoring and coordinating the code agents. In order to support the development of complex multimodal medical artificial intelligence, MARLA has built a hierarchical medical research loop that decomposes the overall goal into multiple specific task sub-loops, and models their dependencies through a DAG, allowing downstream tasks to inherit reusable knowledge, intermediate artifacts, and validated solutions produced by upstream tasks. Experimental results across diverse multimodal medical AI tasks have demonstrated the effectiveness of MARLA in different clinical fields, prediction tasks, and research environments. The results indicate that successful medical artificial intelligence automation requires not only code generation and execution, but also effective research loop construction, continuous supervision, and accumulated research experience. By monitoring progress, diagnosing faults, and improving or restarting sub-loops as necessary, MARLA is able to achieve complex research objectives in a structured and manageable manner. In addition, MARLA continuously improves its cross project research capabilities by converting execution tracking and validation results into reusable memory, skills, tools, and code templates. We hope this work represents a step toward lowering the barriers to medical AI development, enabling clinicians to translate clinical research intent into executable research programs while continually improving future research loops through accumulated experience.

## Data Availability

All data produced are available online.

https://portal.gdc.cancer.gov/projects/TCGA-KIRC

https://www.cancerimagingarchive.net/collection/tcga-kirc/

https://adni.loni.usc.edu/data-samples/access-data/

https://physionet.org/content/mimiciv/

https://physionet.org/content/mimic-cxr-jpg/

https://physionet.org/content/mimic-iv-note/

https://physionet.org/content/mimic-iv-ecg/

## Footnotes

1 https://docs.anthropic.com/en/docs/claude-code

